# Heterologous inactivated virus/mRNA vaccination response to BF.7, BQ.1.1, and XBB.1

**DOI:** 10.1101/2023.02.02.23285205

**Authors:** Fanglei Zuo, Rui Sun, Hassan Abolhassani, Likun Du, Yating Wang, Stelios Vlachiotis, Federico Bertoglio, Maren Schubert, Nima Rezaei, Zahra Chavoshzadeh, Concetta Guerra, Andrea Cavalli, Juni Andréll, Makiko Kumagai-Braesch, Yintong Xue, Yunlong Cao, Michael Hust, Davide F. Robbiani, Xiaoliang Sunney Xie, Lennart Hammarström, Harold Marcotte, Qiang Pan-Hammarström

## Abstract

The emergence of highly immune-escape Omicron variants of the severe acute respiratory syndrome coronavirus 2 (SARS-CoV-2), such as BQ and XBB, has led to concerns about the efficacy of vaccines. Using lentivirus-based pseudovirus neutralizing assay, we showed that heterologous vaccination involving parental mRNA vaccine as a booster or second booster in individuals that received two or three doses of inactivated vaccines strongly augments the neutralizing activity against emerging Omicron subvariants, including BF.7, BQ.1.1, and XBB.1, by 4.3-to 219-folds. Therefore, a heterologous boosting strategy with mRNA-based vaccines should be considered in populations where inactivated vaccines were primarily used.

## Text

The emergence of highly immune-escape Omicron variants of the severe acute respiratory syndrome coronavirus 2 (SARS-CoV-2) has led to concerns about the efficacy of vaccines and therapeutic monoclonal antibodies. Omicron subvariants BA.4.6, BF.7, BQ.1.1, XBB, and XBB.1 harbor the spike protein R346T substitution which contributes to evasion of class III anti-spike monoclonal antibody recognition while XBB, and XBB.1.1 subvariants harbor the F486S substitution which reduces binding for class I and II monoclonal antibodies.^1^ BQ.1, BQ.1.1, XBB, and XBB.1 are increasing rapidly in the United States, India, Europe, and other parts of the world, whereas BF.7 is one of the dominant strains currently circulating in China. Due to humoral immune imprinting, a phenomenon in which initial exposure to the original strain of SARS-CoV-2, by infection or vaccination, limits a person’s future immune response against variants, the bivalent vaccine booster and hybrid immunity may not provide sufficient protection against emerging Omicron subvariants.^2-4^ We have previously shown that an mRNA vaccine booster in individuals vaccinated with two doses of inactivated vaccine significantly increased the level of plasma neutralizing antibodies against Omicron BA.1.^5^ Whether this vaccination strategy retains neutralizing activity against the emerging Omicron subvariants remains unknown.

We used enzyme-linked immunosorbent assay and lentivirus-based pseudovirus neutralizing assay (see the Supplementary Methods section in the Supplementary Appendix), to evaluate the levels of binding and neutralizing antibodies in 77 plasma samples collected from 67 healthy volunteers within 1-5 months after vaccination or infection (Table S1). The participants were grouped based on their vaccination and infection history: three doses of inactivated vaccine; three doses of mRNA vaccine; two doses of inactivated vaccine followed by an mRNA vaccine booster; three doses of inactivated vaccine followed by one or two doses of mRNA vaccine booster; infected during the G614 wave and sampled after one or two subsequent doses of mRNA vaccine; and breakthrough infection during the Omicron BA.1 wave.

In all six groups, the level of receptor-binding domain (RBD)-specific IgG and the neutralizing activity were lower against all Omicron subvariants tested than the original G614 strain, with the lowest level measured against the XBB.1 subvariant (Fig. S1, Fig. 1, and Fig. S2). The level of specific IgG and half-maximal pseudovirus neutralization titers (NT50) against Omicron subvariants in the heterologous vaccination groups were significantly higher than that in individuals receiving three doses of a homologous inactivated vaccine, reaching a level similar to those who received three doses of homologous mRNA-vaccine or a boost of mRNA vaccine after infection or experienced breakthrough infection (Fig. S1 and Fig. 1). The NT50 of heterologous vaccination groups were 155-to 158-folds higher against G614 and 4.3-to 219-folds higher against Omicron subvariants than that of three doses of a homologous inactivated vaccine group (Fig. 1). Although the median NT50 values of each vaccinated/infected group against BF.7, BQ.1.1, and XBB.1 were 4-to 148-fold lower compared to that against G614, the median NT50 values against these Omicron subvariants were 17 to 512 in the heterologous vaccination groups as well as homologous mRNA vaccine group, while those in three doses of a homologous inactivated vaccine group were below the limit of detection (Fig. S2B) and may account for the surge of COVID-19 cases in China where inactivated vaccines are mainly used.

**Figure 1.**
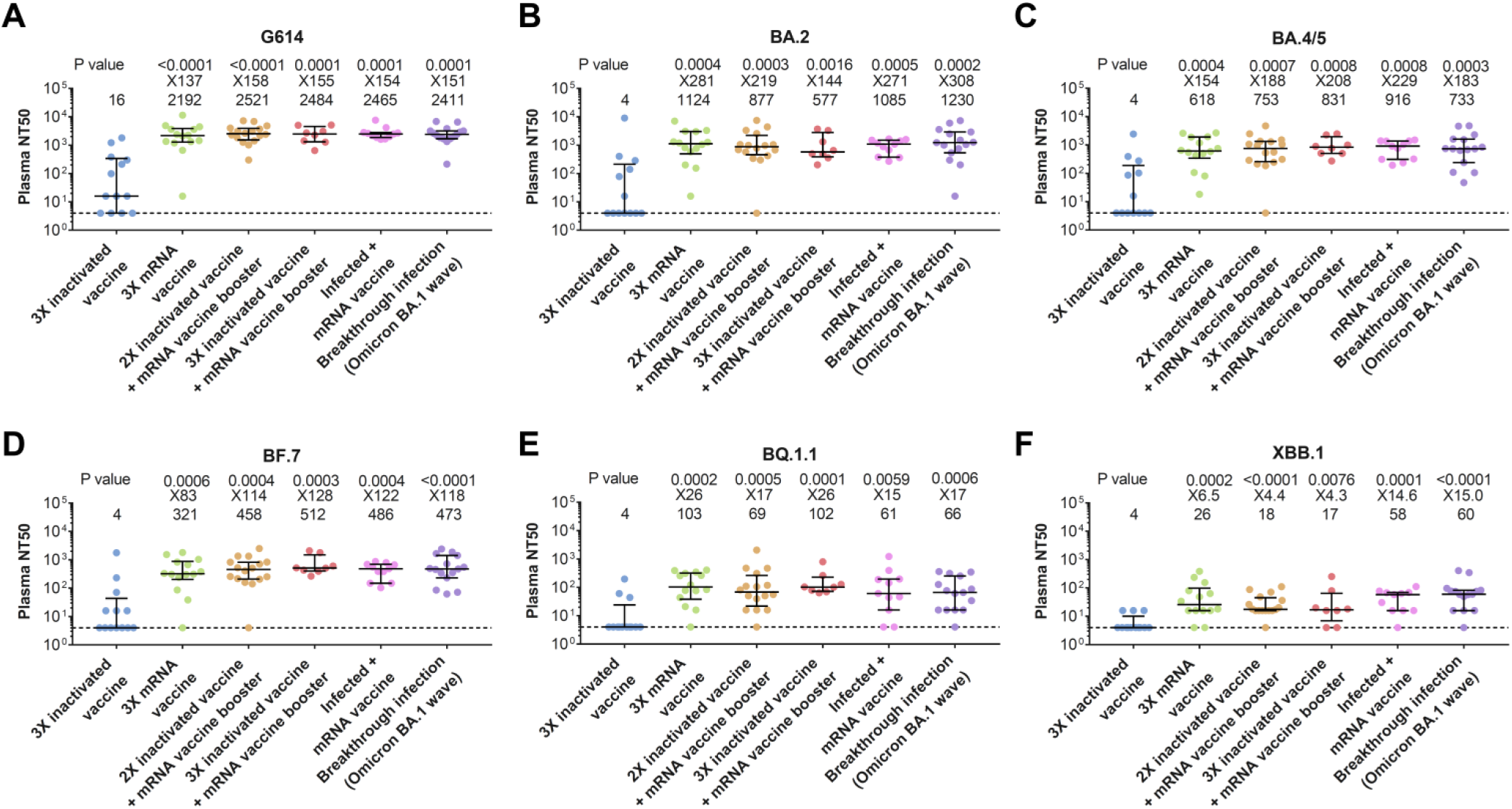
Plasma neutralization activity against G614 and Omicron subvariants. Plasma neutralization activity against the G614 strain of severe acute respiratory syndrome coronavirus 2 (SARS-CoV-2) (panel A) and the Omicron subvariants BA.2 (panel B), BA.4/5 (panel C), BF.7 (panel D), BQ.1.1 (panel E), and XBB.1 (panel F) in participants who received three doses (3X) of inactivated vaccine (n = 13), three doses of mRNA vaccine (n = 14), two doses (2X) of inactivated vaccine followed by an mRNA vaccine booster (n = 16), three doses of inactivated vaccine followed by one or two doses of mRNA vaccine booster (n = 5; 8 samples), one or two doses of mRNA vaccine after infection (G614 wave) (Infected plus mRNA vaccine, n = 9; 11 samples) or had experienced breakthrough infection during the Omicron BA.1 wave (Breakthrough infection, n = 15). The median half-maximal neutralization titers (NT50) and the ratio of the NT50 titer of each group versus the group who received three doses of inactivated vaccine are shown on the top of each panel. Symbols represent individual samples and horizontal black lines indicate the median. Whiskers indicate the interquartile range. A two-sided Mann‒Whitney U test was used to compare the neutralizing activity between three doses of inactivated vaccine group and other groups in each panel. The dashed line indicates the limit of detection.

Thus, among all omicron subvariants tested, the lowest neutralization activity elicited by vaccines and BTI was against BQ.1.1, and XBB.1. Our results suggest that heterologous vaccination involving parental mRNA vaccine as a booster or second booster in individuals that received two/three doses of inactivated vaccines strongly augments the neutralizing activity and may still be effective against emerging Omicron subvariants. Therefore, a heterologous boosting strategy with mRNA-based vaccines should be considered in populations where inactivated vaccines were primarily used.

## Data Availability

The data that support the findings of this study are available within the Article. All other data are available from the corresponding author upon reasonable request.

## Acknowledgments

This work was supported by The European Union’s Horizon 2020 research and innovation program (ATAC, 101003650, D.F.R., M.H., L.H., H.M., Q.P.H), the Center for Innovative Medicine at the Karolinska Institutet (FoUI-963219, Q.P.H), the Swedish Research Council (2019-01302, 2020-06116, Q.P.H), the Knut and Alice Wallenberg Foundation (KAW2020.0102, L.H., Q.P.H), and the Magnus Bergvalls Stiftelse (2022-111, F.Z).

## Supplementary Appendix

## Methods

### Study design

Study inclusion criteria included subjects being above 18 years of age who have received inactivated and/or mRNA vaccines with a documented vaccination history (type of vaccine, number of doses, the interval between the doses, days after the latest dose, if they have been infected), and who were willing and able to provide written informed consent. The study included 77 samples from 67 healthy volunteers (64% females, median age of 31 years) during 2021-2022 who received three doses of inactivated vaccine CoronaVac (Sinovac) or BBIBP-CorV (Sinopharm), or three doses of an mRNA vaccine BNT162b2 (Pfizer–BioNTech) or mRNA-1273 (Moderna), or a combination of both (two doses of inactivated vaccine followed by one or two doses of heterologous mRNA boost; or three doses of inactivated vaccine followed by one or two doses of heterologous mRNA boost), some of whom had experienced breakthrough infections during the Omicron BA.1 wave. Samples were collected 9-144 days (median day 48) after dose three of inactivated vaccine, 11-121 days (median day 28.5) after each mRNA vaccine dose including after mRNA heterologous boost, and 11-43 days (median day 23) after a breakthrough infection (Supplementary Appendix Table 1). Infection was confirmed when an individual tested positive for antigen or qPCR test. Plasma samples from pre-vaccinated, non-infected healthy donors from our cohort (n = 12, Supplementary Appendix Table 1) were also collected as negative controls and used to calculate the cutoff value of the ELISA method. The study was approved by the ethics committees in institutional review board (IRB) of Stockholm, and the Tehran University of Medical Sciences.

### Production of SARS-CoV-2 RBD protein

The RBDs of G614 and Omicron (BA.1, BA.2, BA.4/5) variants were ordered as GeneString from GeneArt (Thermo Fisher Scientific). All sequences of the RBD (aa 319-541 in GenBank: MN908947) were inserted into an *Nco*I/*Not*I compatible variant of an OpiE2 expression vector carrying the N-terminal signal peptide of the mouse Ig heavy chain and a C-terminal 6×His-tag. RBD of G614 and Omicron were expressed in a baculovirus-free expression system in High Five insect cells and purified on HisTrap Excel columns (Cytiva) followed by size-exclusion chromatography on 16/600 Superdex 200-pg columns (Cytiva).^1,2^ The RBDs of Omicron BQ.1.1 and XBB/XBB.1 were ordered from Sino Biologicals.

### Detection of antibodies specific to SARS-CoV-2

For assessing the anti-RBD IgG binding activity, high-binding Corning Half area plates (Corning #3690) were coated overnight at 4°C with RBD derived from the G614 and Omicron BA.1, BA.2, BA.4/5, BQ.1.1, and XBB/XBB.1 (1.7 μg/ml) variants in PBS. Serial dilutions of plasma in 0.1% BSA in PBS were added and plates were subsequently incubated for 1.5 h at room temperature. Plates were then washed and incubated for 1 h at room temperature with horseradish peroxidase (HRP)-conjugated goat anti-human IgG (Invitrogen #A18805) (diluted 1:15 000 in 0.1% BSA-PBS). Bound antibodies were detected using tetramethylbenzidine substrate (Sigma #T0440). The color reaction was stopped with 0.5M H_2_SO_4_ after 10 min incubation and the absorbance was measured at 450 nm using an ELISA plate reader. For each sample, the EC_50_ values were calculated using GraphPad Prism 7.04 software and expressed as relative potency towards an internal calibrant for which the Binding Antibody Unit (BAU) was calculated using the WHO International Standard 20/136 in relation to the G614 RBD. The positive cutoff was calculated as 2 standard deviations (2SD) above the mean of a pool of pre-vaccination samples (n = 12).

### Neutralization assay against pseudotyped SARS-CoV-2

The human-codon optimized gene coding for the S protein of G614, BA.2, and BA.4/5 lacking the C-terminal 19 codons (SΔ19) was synthesized by GenScript. The SΔ19 gene of BF.1, BQ.1.1, and XBB.1 were constructed by site-directed mutagenesis (QuikChange Multi Site-Directed Mutagenesis Kit, Agilent) using BA.2 or BA.4/5 SΔ19 gene as a template. To generate (HIV-1/NanoLuc2AEGFP)-SARS-CoV-2 particles, three plasmids were used, with a reporter vector (pCCNanoLuc2AEGFP), HIV-1 structural/regulatory proteins (pHIVNLGagPol) and SARS-CoV-2 SΔ19 carried by separate plasmids as previously described.^3^ 293FT cells were transfected with 7 µg of pHIVNLGagPol, 7 µg of pCCNanoLuc2AEGFP, and 2.5 µg of a pSARS-CoV-2-SΔ19 carrying the SΔ19 gene from G614 or Omicron variants (at a molar plasmid ratio of 1:1:0.45) using 66 µl of 1 mg/ml polyethylenimine (PEI).

Five-fold serially diluted plasma samples were incubated with pseudotyped SARS-CoV-2 virus (G614, BA.2, BA.4/5, BF.7, BQ.1.1, and XBB.1) for 1 h at 37 °C. The mixture was subsequently incubated with ACE2-expressing HEK293T cells for analyses of G614 or Omicron pseudoviruses for 48 h, after which the cells were washed with PBS and lysed with Luciferase Cell Culture Lysis reagent (Promega). NanoLuc luciferase activity in the lysates was measured using the Nano-Glo Luciferase Assay System (Promega) with a Tecan Infinite microplate reader. The relative luminescence units were normalized to those derived from cells infected with the pseudotyped virus in the absence of plasma samples. The NT50 values for plasma were determined using four-parameter nonlinear regression (the least squares regression method without weighting) (GraphPad Prism 7.04 software).

### Quantification and statistical analysis

Microsoft Excel 2017 was used for data collection for this study. Two-sided Mann‒Whitney U test was performed for comparisons of anti-SARS-CoV-2 binding and neutralizing antibody levels between groups. A Wilcoxon matched-pairs signed rank test was used for the comparison of paired samples. All analyses and data plotting were performed with GraphPad 7.05. A Chi-square test for trend was used to compare the gender between groups. A Kruskal-Wallis analysis of variance (ANOVA) was performed to compare the age and sampling day after vaccination/infection between groups. A p-value less than 0.05 was considered to be statistically significant.

---

**Table S1.** Demographic data of vaccinated individuals.

| Groups | Number of individuals | Number of samples | Male/Female | Median age (IQR)<sup>a</sup>, years | Median sampling day after vaccination/infection (IQR) |
| --- | --- | --- | --- | --- | --- |
| Before vaccination (used for cutoff calculation) | 12 | 12 | 5/7 | 38 (28-48) |  |
| 3 doses of inactivated vaccine | 13 | 13 | 6/7 | 31 (27-56) | 48 (12-92) |
| 3 doses of mRNA vaccine | 14 | 14 | 3/11 | 36 (30-53) | 19 (15-29) |
| 2 doses of inactivated vaccine plus an mRNA vaccine booster | 16 | 16 | 6/10 | 28 (26-29) | 28 (19-39) |
| 3 doses of inactivated vaccine plus mRNA vaccine booster | 5 | 8 (4 after the first dose of mRNA vaccine and 4 after the second dose of mRNA vaccine) | 1/4 | 28 (27-32) | 31 (19-82) |
| Infected plus mRNA vaccine | 9 | 11 (7 after the first dose of mRNA vaccine and 4 after the second dose of mRNA vaccine) | 5/4 | 37 (31-52) | 31 (18-37) |
| Breakthrough infection<sup>b</sup> | 15 | 15 | 4/11 | 34 (27-53) | 23 (17-34) |
| Comparison of groups |  |  | Non-significant<sup>c</sup> | Non-significant<sup>d</sup> | Non-significant<sup>d</sup> |

<sup>a</sup> IQR: Interquartile range.
<sup>b</sup> After two or three doses of either inactivated or mRNA vaccine or a combination of both during the Omicron BA.1 wave.
<sup>c</sup> Chi-square test for trend.
<sup>d</sup> Kruskal-Wallis ANOVA.

**Figure S1.**
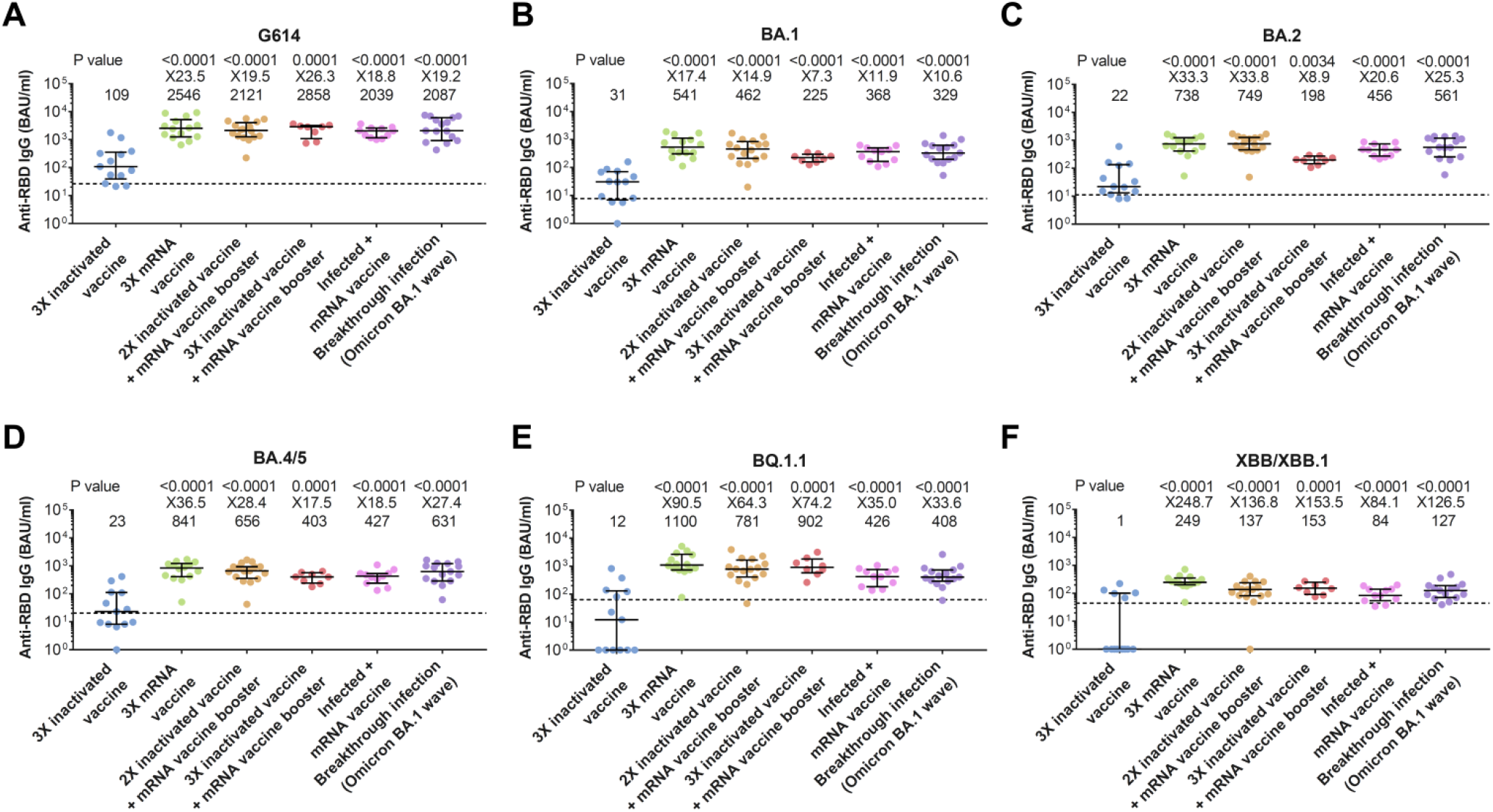
Level of specific IgG antibodies against the receptor binding domain (RBD) of the G614 strain and Omicron subvariants. Level of specific IgG antibodies against the RBD of the G614 strain of severe acute respiratory syndrome coronavirus 2 (SARS-CoV-2) (panel A) and the Omicron subvariants BA.1 (panel B), BA.2 (panel C), BA.4/5 (panel D), BQ.1.1 (panel E), and XBB.1 (panel F) in participants who received three doses (3X) of inactivated vaccine, three doses of mRNA vaccine, two doses (2X) of inactivated vaccine followed by an mRNA vaccine booster, three doses of inactivated vaccine followed by one or two doses of mRNA vaccine booster, one or two doses of mRNA vaccine after infection (G614 wave) (Infected plus mRNA vaccine) or had experienced breakthrough infection during the Omicron BA.1 wave (Breakthrough infection). The median antibody levels and the ratio of the antibody levels of each group versus the group who received three doses of inactivated vaccine are shown on the top of each panel. Symbols represent individual samples and horizontal black lines indicate the median. Whiskers indicate the interquartile range. A two-sided Mann‒Whitney U test was used to compare the antibody levels between three doses (3X) of inactivated vaccine group and other groups in each panel. Antibody levels are presented as binding antibody units (BAU)/ml. The cutoff value (dashed line) is calculated separately for each variant from samples of 12 pre-vaccinated, non-infected individuals.

**Figure S2.**
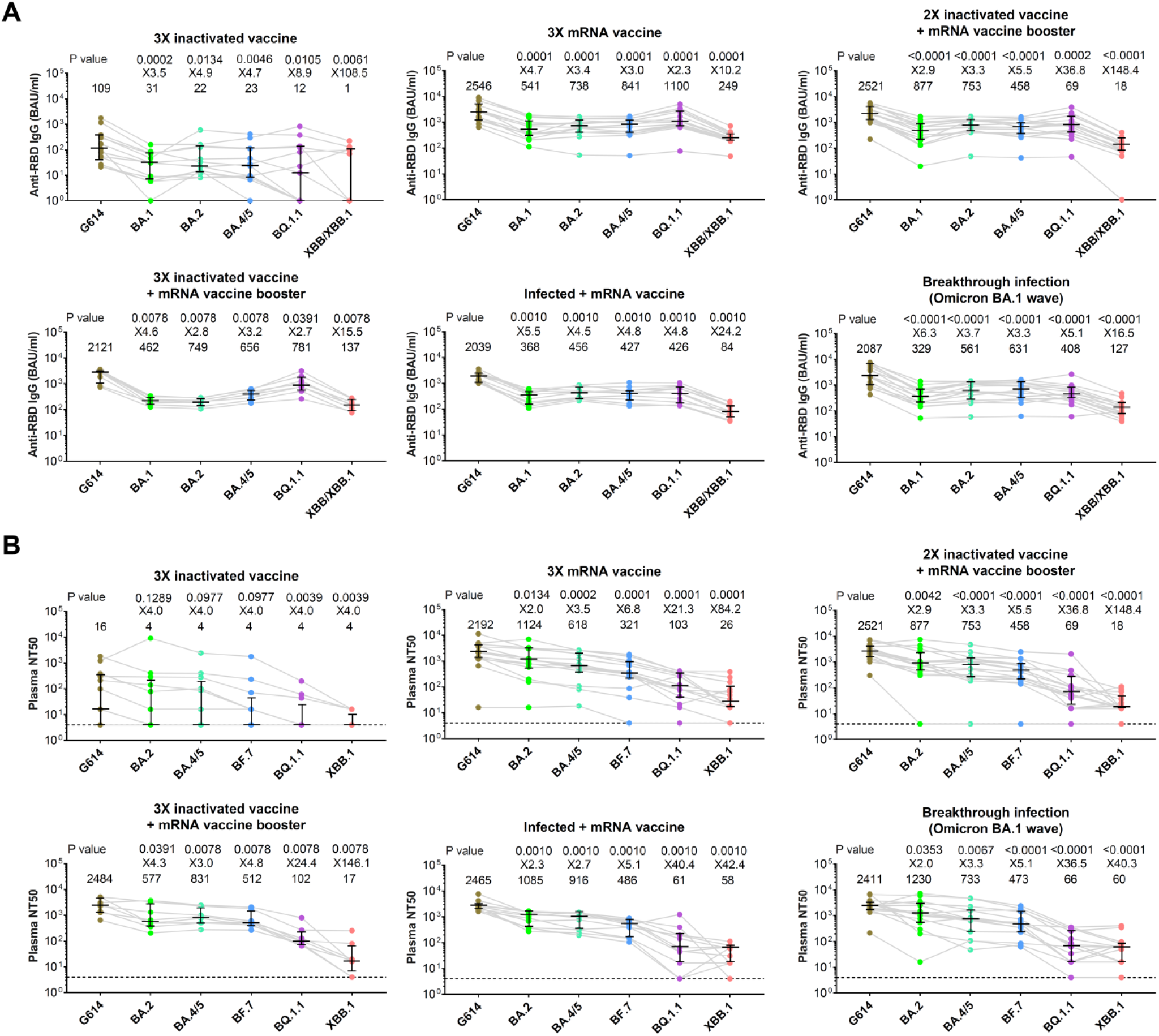
Level of receptor binding domain (RBD)-specific IgG antibodies (A) and neutralization activity (B) against G614 strain and Omicron subvariants. Level of RBD-specific IgG antibodies and half-maximal neutralization titers (NT50) against the G614 strain of SARS-CoV-2 and the Omicron subvariants BA.1, BA.2, BA.4/5, BQ.1.1, and XBB/XBB.1 in participants who received three doses (3X) of inactivated vaccine, three doses of mRNA vaccine, two doses (2X) of inactivated vaccine followed by an mRNA vaccine booster, three doses of inactivated vaccine followed by one or two doses of mRNA vaccine booster, one or two doses of mRNA vaccine after infection (G614 wave) (Infected plus mRNA vaccine) or had experienced breakthrough infection during the Omicron BA.1 wave (Breakthrough infection). The median antibody levels or neutralization titer and the ratio of the antibody levels or neutralization titer against G614 strain to that against each Omicron subvariant are shown on the top of each panel. Antibody levels are presented as binding antibody units (BAU)/ml. Symbols represent individual samples and horizontal black lines indicate the median. Whiskers indicate the interquartile range. The connecting lines between the variants represent matched serum samples. A Wilcoxon matched-pairs signed rank test was used to compare the antibody levels or neutralization titers against G614 strain to that against each Omicron subvariant in each panel. The dashed line indicates the limit of detection.

